# HIV-1 subtype-specific drug resistance on dolutegravir-based antiretroviral therapy: protocol for a multicentre longitudinal study (DTG RESIST)

**DOI:** 10.1101/2024.05.23.24307850

**Authors:** Matthias Egger, Mamatha Sauermann, Tom Loosli, Stefanie Hossmann, Selma Riedo, Niko Beerenwinkel, Antoine Jaquet, Albert Minga, Jeremy L. Ross, Jennifer Giandhari, Roger Kouyos, Richard Lessells

## Abstract

**Introduction:** HIV drug resistance poses a challenge to the United Nation’s goal of ending the HIV/AIDS epidemic. The integrase strand transfer inhibitor (InSTI) dolutegravir, which has a higher resistance barrier, was endorsed by the World Health Organization in 2019 for first-, second-, and third-line antiretroviral therapy (ART). This multiplicity of roles of dolutegravir in ART may facilitate the emergence of dolutegravir resistance.

**Methods and analysis:** DTG RESIST is a multicentre longitudinal study of adults and adolescents living with HIV in sub-Saharan Africa, Asia, and South and Central America who experienced virologic failure on dolutegravir-based ART. At the time of virologic failure whole blood will be collected and processed to prepare plasma or dried blood spots. Laboratories in Durban, Mexico City and Bangkok will perform genotyping. Analyses will focus on (i) individuals who experienced virologic failure on dolutegravir, and (ii) on those who started or switched to such a regimen and were at risk of virologic failure. For population (i), the outcome will be any InSTI drug resistance mutations, and for population (ii) virologic failure defined as a viral load >1000 copies/mL. Phenotypic testing will focus on non-B subtype viruses with major InSTI resistance mutations. Bayesian evolutionary models will explore and predict treatment failure genotypes. The study will have intermediate statistical power to detect differences in resistance mutation prevalence between major HIV-1 subtypes; ample power to identify risk factors for virologic failure and limited power for analysing factors associated with individual InSTI drug resistance mutations.

**Ethics and dissemination:** The research protocol was approved by the Biomedical Research Ethics Committee at the University of KwaZulu-Natal, South Africa, and the Ethics Committee of the Canton of Bern, Switzerland. All sites participate in IeDEA and have obtained ethics approval from their local ethics committee to conduct the additional data collection.

**Registration:** NCT06285110

**Strengths and limitations of this study:**

⍰ DTG RESIST is a large international study to prospectively examine emergent dolutegravir resistance in diverse settings characterised by different HIV-1 subtypes, provision of ART, and guidelines on resistance testing.
⍰ Embedded within the International epidemiology Databases to Evaluate AIDS (IeDEA), DTG RESIST will benefit from harmonized clinical data across participating sites and expertise in clinical, epidemiological, biological, and computational fields.
⍰ Procedures for sequencing and assembling genomes from different HIV-1 strains will be developed at the heart of the HIV epidemic, by the KwaZulu-Natal Research Innovation and Sequencing Platform (KRISP), in Durban, South Africa. Phenotypic testing, Genome Wide Association Study (GWAS) methods and Bayesian evolutionary models will explore and predict treatment failure genotypes.
⍰ A significant limitation is the absence of genotypic resistance data from participants before they started dolutegravir treatment, as collecting and bio-banking pre-treatment samples was not feasible at most IeDEA sites. Consistent and harmonized data on adherence to treatment are also lacking.
⍰ The distribution of HIV-1 subtypes across different sites is uncertain, which may limit the statistical power of the study in analysing patterns and risk factors for dolutegravir resistance. The results from GWAS and Bayesian modelling analyses will be preliminary and hypothesis-generating.

## Introduction

The emergence of HIV drug resistance threatens the United Nations’ goal of ending the HIV/AIDS epidemic by 2030.[1–3] HIV drug resistance has increased substantially with the expansion of access to antiretroviral therapy (ART) in low- and middle-income countries.[4,5] The regimen used widely in recent years combined the nucleoside reverse transcriptase inhibitors (NRTIs) tenofovir and emtricitabine with the non-nucleoside reverse transcriptase inhibitor (NNRTI) efavirenz and thus contained three drugs with a low genetic barrier to resistance. As a result, acquired dual-class NRTI and NNRTI resistance was detected in many people living with HIV (PWH) who experienced virologic failure on this regimen.[4] There has also been a steady increase in pre-treatment drug resistance (PDR). By 2016, NNRTI PDR exceeded 10% in Southern and Eastern Africa and approached this level in Latin America and the Caribbean. The 10% NNRTI PDR threshold is the point at which the World Health Organization (WHO) recommends a change in the standard first-line ART regimen.[6,7]

As a response to the high levels of resistance, WHO endorsed the integrase strand transfer inhibitor (InSTI) dolutegravir in 2019 as the primary choice for both first-line and second-line ART across all groups, including pregnant women and those who may become pregnant.[8] By mid-2022, over 100 countries had incorporated dolutegravir into their treatment guidelines.[9] Dolutegravir has a high genetic barrier to resistance[10,11], and thus far, only few people with HIV (PWH) were found to develop resistance to it.[12–14] The types of mutations that cause dolutegravir resistance may vary by treatment history and HIV-1 subtype. For example, the Arg263Lys mutation appears to be relatively more common in PWH who had not previously received first-generation InSTIs compared to those previously exposed to INSTIs.[15] The Gln148His/Lys/Arg mutation may be less common in subtype C than in other subtypes.[16]

The risk factors and mutation patterns contributing to dolutegravir resistance are not as well-defined as those for older antiretroviral drugs. Dolutegravir’s extensive use in areas with limited resources, where treatment options are more uniform, the reuse of drugs is common, and access to adherence support, viral load, and resistance testing may be limited, could facilitate the emergence of resistance mutations. The DTG RESIST study aims to document the resistance patterns and contexts in which dolutegravir resistance emerges and thus to contribute to safeguarding the long-term sustainability of the global HIV response. DTG RESIST has two parts: the first, a collaborative analysis of existing data from HIV cohort studies, has recently been published.[15] Here, we describe the second part of the study, which prospectively recruits PWH who experience virological failure on dolutegravir-based ART in 17 countries in sub-Saharan Africa, South and Central America, and Asia.

### Study aims and hypotheses

The prospective part of DTG RESIST has three aims:

⍰ In Aim 1, we examine viral sequences to determine the prevalence and patterns of dolutegravir resistance in adults and adolescents living with HIV-1.
⍰ In Aim 2, we combine the HIV drug resistance data from Aim 1 with clinical and programmatic data collected within the IeDEA network to explore the factors associated with emergent dolutegravir resistance.
⍰ In Aim 3, we characterize the phenotypic effect of novel drug resistance mutations or novel combinations of mutations, and explore the mutational pathways of dolutegravir resistance across HIV-1 subtypes.

We hypothesize that the prevalence and spectrum of InSTI drug resistance mutations will differ by HIV-1 subtype and by treatment context (Aim 1); that NRTI resistance, previous exposure to InSTI drugs, male sex, younger age, second- or third-line ART or advanced disease, exposure to rifampicin, and lack of routine viral load monitoring are risk factors for virologic failure and the development of dolutegravir resistance (Aim 2); and that HIV genotypes from patients experiencing failure of dolutegravir-based ART are associated with phenotypic resistance. We further hypothesize that these mutations develop along pathways specific to each HIV-1 subtype, and in combination may compensate for the fitness costs of resistance mutations, i.e., restore replication capacity compared to single mutations (Aim 3).

## Methods and analysis

DTG RESIST is a multicentre longitudinal study enrolling PWH from sub-Saharan Africa, Asia, and South and Central America. <u>Figure 1</u> summarises the study design. PWH who experience virologic failure in routine care (confirmed or based on a single measurement) are enrolled at a dedicated study visit. At the study visit they provide informed consent and a blood sample, which undergoes sequencing if viral load is above 1000 copies/mL. The DTG RESIST data are linked with the routine data collected within the International epidemiology Databases to Evaluate AIDS (IeDEA).

**Figure 1.**
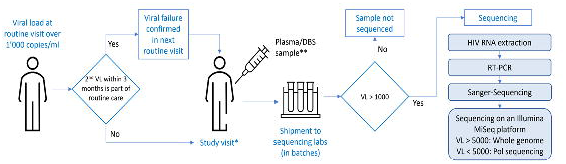
Schematic representation of enrolment, specimen collection and sequencing. VL, viral load; DBS, dried blood spot.

### Setting

DTG RESIST is embedded within IeDEA. Established in 2006 by the National Institute of Allergy and Infectious Diseases (NIAID), IeDEA is a global cohort collaboration that collects HIV/AIDS data from HIV care and treatment programmes. Six of the seven IeDEA regions participate in DTG RESIST: Southern Africa, East Africa, Central Africa, West Africa, Asia Pacific, and the Caribbean, Central and South American region. The IeDEA consortium has been described in detail elsewhere.[17–19] <u>Figure 2</u> shows the countries participating in DTG RESIST and the HIV-1 subtypes and Circulating Recombinant Forms (CRFs) prevalent in the region.

**Figure 2.**
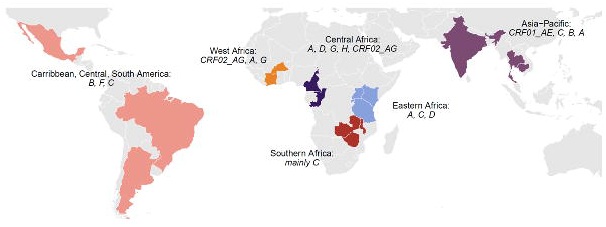
Countries with sites participating in DTG RESIST with the dominant HIV-1 subtypes and circulating recombinant forms (CRFs). The participation of India is pending (awaiting approval). The colours represent the six IeDEA regions.

### Study population, eligibility and recruitment

DTG RESIST enrols adults and adolescents (age 10-17 years) living with HIV who experienced virologic failure during routine care, defined as a single or, if available, a confirmed viral load >1000 copies/mL on any dolutegravir-based combination ART regimen (<u>Figure 2</u>). Eligible PWH were on dolutegravir for at least 3 months and are enrolled consecutively. At enrolment, written informed consent is obtained from the participant or the parents or caretaker, as per local regulations. Key clinical and demographic data are collected using the Research Electronic Data Capture (REDCap) system.[20] The DTG RESIST data will be combined with the clinical and epidemiological data prospectively collected within IeDEA.[17–19]

### Specimen collection and transport

Specimen collection and processing is performed according to WHO/HIVResNet guidance.[21] Whole blood is collected from participants in ethylenediaminetetraacetic acid (EDTA) tubes. Specimens are kept at room temperature (15-30°C) until centrifugation and separation (within 6 hours). All initial processing, centrifugation, pipetting, and aliquoting will be done following standard laboratory bio-safety precautions at a local laboratory equipped to manipulate infectious clinical specimens. Following separation, 4 x 1.5mL plasma cryovials from each participant will be frozen at -80°C. Two dried blood spot (DBS) cards with five spots each are also collected and stored at -20°C. In sites where storage of plasma samples is not possible, only DBS cards will be prepared[22] (<u>Figure 2</u>). A courier service experienced in the transport of clinical specimens ships plasma and DBS specimens on dry ice to the genotyping labs in Durban, Mexico City or Bangkok.

### Sequencing and phenotypic resistance testing

The KwaZulu-Natal Research Innovation and Sequencing Platform (KRISP) in Durban, South Africa [23], the Centro de Investigación en Enfermedades Infecciosas (CIENI), Mexico City[24] and the HIV-NAT virology lab at Chulalongkorn University, Bangkok, Thailand[25] will serve as the regional laboratories. Whenever possible, whole HIV-1 genomes using the Illumina MiSeq Next-Generation Sequencing (NGS) platform will be generated to examine mutations in genetic regions outside pol that could be associated with resistance to InSTIs (<u>Figure 1</u>).[26,27] Alternatively, if low virus loads do not allow whole viral genome sequencing, partial pol sequences including the viral protease, reverse transcriptase, and integrase genes will be generated. We will perform phenotypic testing on about 100 selected samples focussing on non-B subtype viruses with major InSTI resistance mutations,[28] including sequences with complex patterns of drug resistance mutations. Phenotypic resistance testing will be performed using the PhenoSense Integrase assay (Monogram Biosciences, San Francisco, CA[29]) which provides an in-vitro-measurement of the integrase half maximal inhibitory concentration (IC50) to dolutegravir and the viral replication capacity. In order to test the reliability of genotypic prediction, we will compare the IC50 with the genotypic sensitivity scores predicted from the Stanford HIVdb, REGA, or ANRS algorithms.[30] We will use a viral genome wide association study (GWAS) approach to explore the effect of mutations outside of the integrase.

### Outcomes and statistical analyses

We define two analysis populations to address Aim 1 and Aim 2: (i) all individuals who experienced virologic failure on a dolutegravir-based first-, second- or third-line regimen in one of the participating cohorts and (ii) all who started ART on a dolutegravir-based regimen or switched to such a regimen and were at risk of developing virologic failure (the source population for (i)). The outcome of interest in population (i) will be any InSTI drug resistance mutations whereas in (ii) it will be virologic failure as defined above.

We will analyse population (i) using random-intercept logistic regression and population (ii) using parametric survival models[31], stratified by treatment program or country to account for heterogeneity across sites. We will measure time from the date of switching to DTG-based ART (or from six months after initiating DTG-based first-line ART) to the earliest of onset of virologic failure, date of death or date of last follow-up visit. Exposure variables include age, sex, type of dolutegravir-based ART (initiated first-line, switched to first-, second- or third-line), previous exposure to InSTI drugs, pre-existing resistance mutations (from HIV-1 genotypes or predicted by the approach described below), advanced disease (as indexed by CD4 count <200 cells/μL), exposure to rifampicin, a history of ART interruptions, and viral load monitoring coverage and frequency (using approaches used in previous analyses[32,33]). We will follow recommendations on the conduct and reporting of prognostic research, including on the choice of variables for inclusion in models, the handling of continuous variables and of non-linear relationships.[34] Missing values will be imputed using multiple imputation, assuming data are missing at random.[35]

We will explore the use of conjunctive Bayesian network models to assess the mutational pathways of DTG resistance (Aim 3).[36–38] The order of mutation accumulation and interdependence between mutations (i.e., whether the acquisition of one mutation is less or more likely in the presence of other mutations) will be inferred from the co-occurrence patterns of mutations. We will learn these models for each subtype separately to identify subtype-specific mutational pathways. In exploratory analyses, we will search for novel resistance mutations that may mediate treatment failure and assess the impact of previously proposed mutations such as mutations outside the HIV-1 integrase.[39,40] We will combine the genotype data generated in Aim 1 with viral whole genome sequences from phylogenetically matched controls from the Los Alamos HIV sequence database. In this pooled data-set, we will use GWAS methodology (as implemented in the PLINK software[41]) to identify the amino-acid mutations associated with failure.

### Sample size and power calculations

Based on a survey of participating sites and recruitment so far, we estimate that about 1500 PWH will experience virologic failure, including 900 on first-line and 600 patients on second- or third-line ART. The number of individuals with drug resistance mutations will depend on the distribution across groups of patients initiating or switching to different lines of dolutegravir-based ART, the proportion switching with detectable VL, and other factors.

For Aim 1, we will have reasonable power to detect differences between HIV-1 subtypes. For example, comparing subtype C with subtype B and assuming a total sample of 250 in each group, will allow us to detect a difference in the prevalence of resistance mutations of 3% versus 9% with 81% power. The test statistic used for these calculations (done in PASS 15[42]) is the two-sided Z-Test; the significance level of the test is 0.0500. The power will be lower for comparisons with other subtypes with fewer patients. For Aim 2, with 1500 patients, we will have ample power to identify risk factors for virologic failure in univariable and multivariable models. Power will be more limited for analyses of factors associated with the presence of InSTI DRMs. Assuming that 10% of patients on first-line ART, 20% on second-line ART and 30% on third-line ART will develop virologic failure within 2 years, and further assuming that among patients failing first-line ART 1-3% will develop InSTI DRMs, whereas 10-20% of patients failing second-line and 20-30% of patients failing third-line ART will develop InSTI DRMs, we can expect 84 to 157 patients with InSTI DRMs. As shown in <u>Figure 3</u> (from PASS 15[42]), in univariate analyses we will have power of 80% or above to detect risk factors with an odds ratio (OR) of 2 or above for a range of exposure prevalence.[43] Power will be lower in the multivariable models, which should not include more than 5-10 variables.[44]

**Figure 3.**
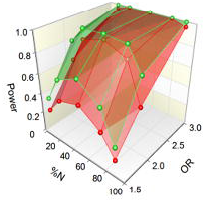
Power vs. exposure prevalence (%N) and odds ratio (OR) for 84 (red) and 157 (green) patients developing InSTI DRMs.

The first PWH was recruited into the study on 13 June 2022 in Kanyama, Zambia. At the time of writing, over 550 PWH had been enrolled.

### Patient and public involvement

There was no involvement of patients or the public in the development of the research questions or the study methods.

## Discussion

In low- and middle-income countries, WHO recommends a public health approach to providing and monitoring antiretroviral therapy,[45–48] including standardized treatment regimens, simplified patient monitoring and clinical decision-making, population-level surveillance, and monitoring of HIV drug resistance.[6,49] WHO guidelines also recommend the use of dolutegravir in first-line, second-line and third-line ART regimens.[8] This multiplicity of roles of dolutegravir in the public health approach to ART combined with limited access to viral load and HIV drug resistance testing, may facilitate the emergence of dolutegravir resistance. DTG RESIST is a timely study addressing this threat.

DTG RESIST leverages the unique resource of IeDEA, a large international research consortium and rich resource for globally diverse HIV/AIDS data.[17–19,50] This brings four key strengths to the study: i) pooling of data across a network of cohorts to explore patterns and determinants of a rare event, dolutegravir resistance; ii) inclusion of multiple HIV-1 subtypes across diverse geographic areas; iii) harmonized clinical and drug resistance data across participating sites; and iv) combined expertise in the clinical, epidemiological, biological, and computational fields required to assess the genotype-phenotype mapping. The strength of pooling data across multiple cohorts and regions is highlighted by the many important collaborative publications of IeDEA.[32,51–57]

DTG RESIST will generate whole HIV-1 genomes and develop robust laboratory procedures to amplify, sequence and assemble the genomes from the diverse set of HIV-1 strains to be included from the multiregional network.[27] The spectrum of dolutegravir-selected mutations and their effects on phenotypic susceptibility in non-B subtypes is yet to be fully characterized. HIV-1 main group (group M) has diversified into nine subtypes (A-D, D-H, J-K), six sub-subtypes (A1-A4, F1-F2), multiple circulating recombinant forms (CRFs) and thousands of unique recombinant forms.[58,59] Most people living with HIV are infected with non-B subtypes but much of the evidence about ART outcomes and HIVDR is based on subtype B viruses from Europe and North America. HIV-1 subtype could influence treatment outcomes and the emergence of drug resistance.[60–62] Finally, phenotypic testing of non-B subtype viruses with major InSTI resistance mutations, and GWAS to explore the effect of mutations outside of the integrase will likely provide new insights into the relevant drug resistance pathways. Bayesian evolutionary models of accumulating mutations will be used to predict the expected genotype at treatment failure.[36]

There are also limitations to the approach. An important weakness is the lack of genotypic resistance data from prior to dolutegravir treatment for most participants. We considered collecting and bio-banking samples from all people starting dolutegravir-based ART and then having paired samples from pre-treatment and at time of virologic failure but this was not deemed feasible at the vast majority of IeDEA sites. We will explore whether these missing baseline genotypes can be imputed by adapting models to predict expected resistance genotypes based on treatment and clinical history. Other challenges include the lack of harmonized data on adherence. There is uncertainty about the distribution of HIV-1 subtypes across sites and the statistical power of the analyses of patterns and risk factors for dolutegravir resistance will be limited. To address these issues, we will consider pooling our data with those from other related studies.[15] The results of the GWAS and Bayesian network analyses may be subject to overinterpretation due to strong model assumptions and limited sample size. To avoid this, we will qualify these as preliminary and hypothesis-generating studies requiring validation.

## Conclusions

Monitoring the emergence of dolutegravir resistance is essential to prevent resistance at the individual and the population level and to ensure the long-term sustainability of ART. DTG RESIST will make an important contribution to a better understanding of the epidemiology and biology of dolutegravir resistance across a wide range of settings.

## Data Availability

All data produced in the present study are available upon reasonable request to the authors

## Ethics and dissemination

The research protocol was approved by the Biomedical Research Ethics Committee at the University of KwaZulu-Natal and the Ethics Committee of the Canton of Bern. All sites participate in IeDEA and have approval from their Institutional Review Boards (IRB) or Ethics Committees to contribute data to this research network. In addition, each participating site obtained ethics approval from their local ethics committee to conduct the specific additional data collection, including determination of adequate provisions solicitation assent by adolescents or parental consent. The research findings will be shared through open access publications and in dissemination meetings with local stakeholders, healthcare providers and communities.

## Acknowledgments

We thank all the people living with HIV who contribute data to this study and all collaborators at participating sites.

## Footnotes

### Twitter

@eggersnsf, @rjlessels

### Contributors

The study concept and design was conceived by ME, NB, RK, SH and RL. MS, TL, SR are responsible for data management and data quality. JG and RK will supervise laboratory work. Analyses will be conducted by TL, NB, RK, and ME. ME prepared the first draft of the manuscript. All authors critically revised the manuscript and approved the submitted version.

### Patient and public involvement

Patients and/or the public were not involved in the design, or conduct, or reporting, or dissemination plans of this research.

### Data sharing

De-identified participant data and a data dictionary will be made available and shared under a data transfer agreement. Requests for access to DTG RESIST data should be sent to. Nucleotide sequences will be made available on GenBank where local regulations allow data sharing.

### Provenance and peer review

Not commissioned; externally peer reviewed.

## References

1 Hamers RL, Rinke de Wit TF, Holmes CB. HIV drug resistance in low-income and middle-income countries. Lancet HIV. 2018;5:e588–96.

2 World Health Organization. WHO HIV drug resistance report 2017. Geneva: World Health Organization 2017.

3 World Health Organization. WHO HIV drug resistance report 2019. Geneva: World Health Organization 2019.

4 Gregson J, Tang M, Ndembi N, et al. Global epidemiology of drug resistance after failure of WHO recommended first-line regimens for adult HIV-1 infection: a multicentre retrospective cohort study. The Lancet Infectious Diseases. 2016;16:565–75.

5 Gupta RK, Gregson J, Parkin N, et al. HIV-1 drug resistance before initiation or re-initiation of first-line antiretroviral therapy in low-income and middle-income countries: a systematic review and meta-regression analysis. The Lancet Infectious Diseases. 2018;18:346–55.

6 World Health Organization. WHO Global action plan on HIV drug resistance 2017–2021.Geneva: World Health Organization 2017.

7 World Health Organization. WHO Guidelines on the public health response to pretreatment HIV drug resistance. Supplement to the 2016 consolidated guidelines on the use of antiretroviral drugs for treating and preventing HIV infection: second edition June 2016. Geneva: World Health Organization 2017.

8 WHO. Update of recommendations on first- and second-line antiretroviral regimens. Geneva, Switzerland: World Health Organization; WHO. 2019;3.

9 WHO Global HIV Programme. WHO HIV Policy Adoption and Implementation Status in Countries. Geneva, Switzerland: World Health Organizaiton 2022.

10 Llibre JM, Pulido F, García F, et al. Genetic barrier to resistance for dolutegravir. AIDS reviews. 2015;17:56–64.

11 Cottrell ML, Hadzic T, Kashuba ADM. Clinical pharmacokinetic, pharmacodynamic and drug-interaction profile of the integrase inhibitor dolutegravir. Clinical Pharmacokinetics. 2013;52:981–94.

12 Cevik M, Orkin C, Sax PE. Emergent resistance to dolutegravir among instinaive patients on first-line or second-line antiretroviral therapy: A review of published cases. Open Forum Infectious Diseases. 2020;7. doi: 10.1093/OFID/OFAA202

13 Pena MJ, Chueca N, D’Avolio A, et al. Virological failure in HIV to triple therapy with dolutegravir-based firstline treatment: Rare but possible. Open Forum Infectious Diseases. 2019;6. doi: 10.1093/ofid/ofy332

14 Scherrer AU, Yang W-L, Kouyos RD, et al. Successful Prevention of Transmission of Integrase Resistance in the Swiss HIV Cohort Study. Journal of Infectious Diseases. 2016;214:399–402.

15 Loosli T, Hossmann S, Ingle SM, et al. HIV-1 drug resistance in people on dolutegravir-based antiretroviral therapy: a collaborative cohort analysis. The Lancet HIV. 2023;10:e733–41.

16 Arimide DA, Szojka ZI, Zealiyas K, et al. Pre-Treatment Integrase Inhibitor Resistance and Natural Polymorphisms among HIV-1 Subtype C Infected Patients in Ethiopia. Viruses. 2022;14:729.

17 Chammartin F, Ostinelli CHD, Anastos K, et al. International epidemiology databases to evaluate AIDS (IeDEA) in sub-S-aharan Africa, 2012–2019. BMJ Open. 2020;10:e035246.:10.

18 McGowan CC, Cahn P, Gotuzzo E, et al. Cohort Profile: Caribbean, Central and South America Network for HIV research (CCASAnet) collaboration within the International Epidemiologic Databases to Evaluate AIDS (IeDEA) programme. Int J Epidemiol. 2007;36:969–76.

19 Gange SJ, Kitahata MM, Saag MS, et al. Cohort profile: the North American AIDS Cohort Collaboration on Research and Design (NA-ACCORD). Int J Epidemiol. 2007;36:294–301.

20 Harris PA, Taylor R, Minor BL, et al. The REDCap consortium: Building an international community of software platform partners. J Biomed Inform. 2019;95:103208.

21 Bertagnolio S, Derdelinckx I, Parker M, et al. World Health Organization/HIVResNet Drug Resistance Laboratory Strategy. Antivir Ther (Lond). 2008;13 Suppl 2:49–57.

22 Singh D, Dhummakupt A, Siems L, et al. Alternative Sample Types for HIV-1 Antiretroviral Drug Resistance Testing. J Infect Dis. 2017;216:S834–7.

23 Manasa J, Danaviah S, Pillay S, et al. An affordable HIV-1 drug resistance monitoring method for resource limited settings. J Vis Exp. 2014;85:51242.

24 Ávila-Ríos S, García-Morales C, Matías-Florentino M, et al. Pretreatment HIV-drug resistance in Mexico and its impact on the effectiveness of first-line antiretroviral therapy: a nationally representative 2015 WHO survey. Lancet HIV. 2016;3:e579–91.

25 Sirivichayakul S, Kantor R, DeLong AK, et al. Transmitted HIV drug resistance at the Thai Red Cross anonymous clinic in Bangkok: results from three consecutive years of annual surveillance. J Antimicrob Chemother. 2015;70:1146–9.

26 Derache A, Iwuji CC, Baisley K, et al. Impact of Next-generation Sequencing Defined Human Immunodeficiency Virus Pretreatment Drug Resistance on Virological Outcomes in the ANRS 12249 Treatment-as-Prevention Trial. Clin Infect Dis. 2019;69:207–14.

27 Gall A, Ferns B, Morris C, et al. Universal amplification, next-generation sequencing, and assembly of HIV-1 genomes. J Clin Microbiol. 2012;50:3838–44.

28 Rhee S-Y, Grant PM, Tzou PL, et al. A systematic review of the genetic mechanisms of dolutegravir resistance. J Antimicrob Chemother. 2019;74:3135–49.

29 PhenoSense Integrase | Monogram Biosciences. https://monogrambio.labcorp.com/resources/phenotyping/phenosense-integrase (accessed 19 January 2024)

30 Liu TF, Shafer RW. Web resources for HIV type 1 genotypic-resistance test interpretation. Clin Infect Dis. 2006;42:1608–18.

31 Royston P, Parmar MKB. Flexible parametric proportional-hazards and proportional-odds models for censored survival data, with application to prognostic modelling and estimation of treatment effects. Stat Med. 2002;21:2175–97.

32 Haas AD, Keiser O, Balestre E, et al. Monitoring and switching of first-line antiretroviral therapy in adult treatment cohorts in sub-Saharan Africa: collaborative analysis. Lancet HIV. 2015;2:e271–278.

33 Keiser O, Chi BH, Gsponer T, et al. Outcomes of antiretroviral treatment in programmes with and without routine viral load monitoring in Southern Africa. AIDS. 2011;25:1761–9.

34 Riley RD, Windt D van der, Croft P, et al., editors. Prognosis Research in Healthcare: Concepts, Methods, and Impact. Oxford, New York: Oxford University Press 2019.

35 Sterne JAC, White IR, Carlin JB, et al. Multiple imputation for missing data in epidemiological and clinical research: potential and pitfalls. BMJ. 2009;338:b2393.

36 Beerenwinkel N, Montazeri H, Schuhmacher H, et al. The individualized genetic barrier predicts treatment response in a large cohort of HIV-1 infected patients. PLoS Comput Biol. 2013;9:e1003203.

37 Montazeri H, Günthard HF, Yang W-L, et al. Estimating the dynamics and dependencies of accumulating mutations with applications to HIV drug resistance. Biostatistics. 2015;16:713–26.

38 Montazeri H, Kuipers J, Kouyos R, et al. Large-scale inference of conjunctive Bayesian networks. Bioinformatics. 2016;32:i727–35.

39 Power RA, Davaniah S, Derache A, et al. Genome-Wide Association Study of HIV Whole Genome Sequences Validated using Drug Resistance. PLoS ONE. 2016;11:e0163746.

40 Power RA, Parkhill J, de Oliveira T. Microbial genome-wide association studies: lessons from human GWAS. Nat Rev Genet. 2017;18:41–50.

41 Purcell S, Neale B, Todd-Brown K, et al. PLINK: A Tool Set for Whole-Genome Association and Population-Based Linkage Analyses. The American Journal of Human Genetics. 2007;81:559–75.

42 PASS 15 Power Analysis and Sample Size Software. NCSS, LLC. Kaysville, Utah, USA. 2015. https://ncss.com/software/pass

43 Hsieh FY, Bloch DA, Larsen MD. A simple method of sample size calculation for linear and logistic regression. Stat Med. 1998;17:1623–34.

44 Austin PC, Steyerberg EW. Events per variable (EPV) and the relative performance of different strategies for estimating the out-of-sample validity of logistic regression models. Stat Methods Med Res. 2017;26:796–808.

45 World Health Organization. WHO Update of Recommendations on First- and Second-Line Antiretroviral Regimens. Geneva: World Health Organization 2019.

46 World Health Organization. WHO Consolidated guidelines on the use of antiretroviral drugs for treating and preventing HIV infection. Geneva: World Health Organization 2016.

47 World Health Organization. WHO Updated recommendations on first-line and second-line antiretroviral regimens and post-exposure prophylaxis and recommendations on early infant diagnosis of HIV. Interim guidance. Geneva: World Health Organization 2018.

48 Gilks CF, Crowley S, Ekpini R, et al. The WHO public-health approach to antiretroviral treatment against HIV in resource-limited settings. Lancet. 2006;368:505–10.

49 Bertagnolio S, Beanland RL, Jordan MR, et al. The World Health Organization’s Response to Emerging Human Immunodeficiency Virus Drug Resistance and a Call for Global Action. J Infect Dis. 2017;216:S801–4.

50 Egger M, Ekouevi DK, Williams C, et al. Cohort Profile: the international epidemiological databases to evaluate AIDS (IeDEA) in sub-Saharan Africa. Int J Epidemiol. 2012;41:1256–64.

51 Duda SN, Farr AM, Lindegren ML, et al. Characteristics and comprehensiveness of adult HIV care and treatment programmes in Asia-Pacific, sub-Saharan Africa and the Americas: results of a site assessment conducted by the International epidemiologic Databases to Evaluate AIDS (IeDEA) Collaboration. J Int AIDS Soc. 2014;17:19045.

52 Parcesepe AM, Mugglin C, Nalugoda F, et al. Screening and management of mental health and substance use disorders in HIV treatment settings in low- and middle-income countries within the global IeDEA consortium. Journal of the International AIDS Society. 2018;21:e25101.

53 Tymejczyk O, Brazier E, Yiannoutsos CT, et al. Changes in rapid HIV treatment initiation after national “treat all” policy adoption in 6 sub-Saharan African countries: Regression discontinuity analysis. PLOS Medicine. 2019;16:e1002822.

54 Nash D, Katyal M, Brinkhof MW, et al. Long-term immunologic response to antiretroviral therapy in low-income countries: a collaborative analysis of prospective studies. AIDS. 2008;22:2291–302.

55 Anderegg N, Panayidou K, Abo Y, et al. Global Trends in CD4 Cell Count at the Start of Antiretroviral Therapy: Collaborative Study of Treatment Programs. Clin Infect Dis. 2018;66:893–903.

56 Chi BH, Yiannoutsos CT, Westfall AO, et al. Universal definition of loss to follow-up in HIV treatment programs: a statistical analysis of 111 facilities in Africa, Asia, and Latin America. PLoS Med. 2011;8:e1001111.

57 Zurcher K, Ballif M, Fenner L, et al. Drug susceptibility testing and mortality in patients treated for tuberculosis in high-burden countries: a multicentre cohort study. Lancet Infect Dis. 2019;19:298–307.

58 Hemelaar J. The origin and diversity of the HIV-1 pandemic. Trends in Molecular Medicine. 2012;18:182–92.

59 Hemelaar J. Implications of HIV diversity for the HIV-1 pandemic. J Infect. 2013;66:391–400.

60 Lessells RJ, Katzenstein DK, de Oliveira T. Are subtype differences important in HIV drug resistance? Curr Opin Virol. 2012;2:636–43.

61 Kantor R, Smeaton L, Vardhanabhuti S, et al. Pretreatment HIV Drug Resistance and HIV-1 Subtype C Are Independently Associated With Virologic Failure: Results From the Multinational PEARLS (ACTG A5175) Clinical Trial. Clin Infect Dis. 2015;60:1541–9.

62 Scherrer AU, Ledergerber B, von Wyl V, et al. Improved virological outcome in White patients infected with HIV-1 non-B subtypes compared to subtype B. Clin Infect Dis. 2011;53:1143–52.

